# Universality of Universal Health Coverage: a Scoping Review

**DOI:** 10.1101/2022.05.28.22275496

**Authors:** Aklilu Endalamaw, Charles F Gilks, Fentie Ambaw, Yibeltal Assefa

## Abstract

**Background:** Universal health coverage (UHC) is achieved in the primary healthcare pathways. UHC is about population coverage, services coverage and financial protection. Tracer indicators are used to measure the progress towards UHC. There is inadequate evidence that assess the extent of the universality of UHC. Evidence is, therefore, needed to map the existing literature and summarize the issues covered in the dimensions of UHC.

**Methods:** A systematic search was carried out in the Web of Science and PubMed databases. Hand searches were also conducted to find articles from Google Scholar, the World Bank Library, the World Health Organization Library, the United Nations Digital Library Collections, and Google. Articles on UHC coverage, financial risk protection, quality of care, and inequity were included. A stepwise approach was used to identify and select relevant studies, conduct data charting, collation and summarization, as well as report results. Simple descriptive statistics and narrative synthesis were used to present the findings.

**Results:** Forty-seven papers were included in the final review. One-fourth of the articles (25.5%) were from the African region and 29.8% were from lower-middle-income countries. More than half of the articles (54.1%) used a quantitative research approach. Of included articles, coverage was assessed by 53.2% of articles; financial risk protection by 27.7%, inequity by 25.5% and quality by 6.4% of the articles as their main research objectives or mentioned in result section. Most (42.5%) of articles investigated health promotion and 2.1% palliation and rehabilitation services. Policy and health care level and cross-cutting barriers were identified in the progress of UHC.

**Conclusions:** The results of the study showed that majorities of evidence were from Africa’s region. Methodologically, the quantitative approach was a more frequently used research design to investigate UHC. Palliation and rehabilitation health care services need attention in the analysis of the progress towards UHC. The finding of the current evidence is noteworthy to focus on quality and inequity of health services in the future UHC research. Comprehensive evidence is needed to fully understand and progressively realize UHC.

## Introduction

Universal health coverage (UHC) is a multi-dimensional concept that includes population coverage, services coverage and financial protection as its building blocks, as well as all individuals and communities (equity), quality, and types of health services in its integrated definition (1). UHC is thought to have the ability to improve the population’s health, economic progress, and social justice (2, 3). UHC work is essential to minimize disparities, promote effective and comprehensive health governance, and build resilient health systems (4). Many components of primary health care, such as proper nutrition, clean water and sanitation, maternal and child health care, immunisation, local disease control, accessible treatment, and drug provision, are intertwined with UHC (5).

The United Nation’s (UN) post-2015 goals described UHC as the predominant design for achieving sustainable health goals of 2030 (6). Furthermore, at the UHC 2020 high-level meeting, UHC was declared an urgent priority for addressing global health crises, including the pandemic COVID-19 (7). This could be accomplished by carrying out activities aimed at providing the desired percentage of the population with essential quality healthcare services while keeping healthcare costs to an acceptable minimum. The UN General Assembly declared at its 73rd session that one billion more people would have access to health care by 2023 (8). The UN also agreed to make essential health services available to 80 percent of the population by 2030, with no catastrophic health expenditures (6).

Tracking the progress of UHC at the global and national levels is essential to identify successes and challenges as well as put forward recommendations. The World Health Organization (WHO) launched its recommendation on research for UHC in 2010 and 2013 (9, 10). The World Bank (WB) also did the same in 2013 (11). Following that, WHO and the WB established core tracers of health service coverage, based on the target of sustainable development goal (SDG) 3.8.1, to quantify UHC using a single measurable value, which integrates the majority of its tracers (12). under the main theme of (1) reproductive, maternal, neonatal, and child health (RMNC), (2) infectious diseases (IDs), (3) non-communicable diseases (NCDs), and (4) service capacity and access (SCA), category one indicators include family planning (FP), antenatal care (ANC) and delivery, immunisation coverage, child-care seeking behaviour for pneumonia; category two indicators include tuberculosis (TB) treatment success, antiretroviral therapy (ART) for HIV, bed net for malaria prevention, and water and adequate sanitation; category three indicators include prevention and treatment of high blood pressure, prevention and treatment of high blood glucose, cervical cancer screening, and tobacco (non-) smoking; and category four indicators include basic hospital access, health worker density, access to essential medicines, and health security. SDG 3.8.2 is about financial risk protection, which is typically measured by catastrophic health expenditure (CHE) and the level of the impoverishment caused by health costs (12).

In addition to these internationally agreed tracer indicators to monitor UHC, other global organisations and countries created additional or new effective service coverage tracer indicators. Notably, the Global Burden Disease Collaborators-2019 (GBD-2019) developed twenty-three indicators of UHC effective service coverage (13). Researchers from China developed Chinese indices of accessibility and affordability indicators to assess the overall state of UHC (14).

Several studies have been conducted on UHC all over the world. These studies have utilized varying methods, measurement metrics, and indicators that require mapping and characterization. A scoping review of the studies on UHC and its dimensions is crucial to map and characterize the existing studies and identify the gaps in evidence towards UHC. This will also help to identify key concepts, gaps in the research, and types and sources of evidence to inform practice, policymaking, and research (15). While no prior studies have been conducted to identify and map the available evidence on UHC, other related studies such as “a synthesis of conceptual literature and global debates” (1) and a scoping review of “implementation research approaches of UHC” (16) are available; however, both did not map and synthesize existing UHC researches. In addition to these literatures, another study assessed the hegemonic nature of UHC in health policy and described historical background how UHC emerged, and frequency of UHC mentioned in all fields of articles available in PubMed database (17). Taking in account UHC as the main research topic, none of the previous studies addressed the universality of UHC in terms of its building blocks (population coverage, service coverage, and financial coverage). Furthermore, those studies did not aim to map existing research on UHC and summarize the findings of each study included in the review. The types of health services (health promotion, disease prevention, treatment, rehabilitation, and palliative care) as an integrated definition of UHC, and how much each is covered in UHC studies, also were not taken into account in those studies.

The goals of this scoping review are to first determine the characteristics of UHC studies, their approaches, and the distribution of articles across WHO regions; second, to synthesize UHC research findings; and third, to inform priority research questions for future studies. To the best of our knowledge, this will be the first scoping review describing available research outputs with their findings on UHC, universality of UHC studies in terms of incorporating UHC dimensions, and types of health services covered in its research.

## Methods

### Identifying a research question

The protocol of this scoping review is available elsewhere https://doi.org/10.21203/rs.3.rs-1082468/v1. The overall activities adhered to the Arksey and O’Malley’s (2005) scoping review framework (18), which was expanded with a methodological enhancement for scoping review projects (19), and the Joanna Briggs Institution framework (20). The review followed five steps: (1) identifying research questions, (2) identifying relevant studies, (3) study selection, (4) data charting, and (5) collation, summarization and reporting of results. The checklist Preferred Reporting Items for Systematic reviews and Meta-analysis extension for Scoping Reviews were used (Supplementary file-1) (21).

The research questions were developed by AE in collaboration with YA. The Population, Concept, and Context framework was used to determine the eligibility of research questions. According to the framework, the population consisted of people of any gender at any age, as well as health institutions and the concept was UHC and interrelated objectives in the global context.

### Identifying relevant studies

Web of Science, PubMed and Google Scholar were used to find literature in the field. Hand searches were also used to find articles from Google Scholar, WB Library, WHO Library, UN Digital Library Collections, and Google. Using the relevant keywords and/or phrases, a comprehensive search strategy was established. Universal, health, “health care”, healthcare, “health service”, quality, access, coverage, equity, disparity, inequity, equality, inequality, expenditure, and cost were search words and/or phrases. “AND” or “OR” Boolean operators were used to broaden and narrow the specific search results. Search strings were formed in accordance with the need for databases (supplementary file-2). The articles were imported into EndNote desktop version x7, which was used to perform an automatic duplication check. Manual duplication removal was also performed. The database search strategies are shown in the supplementary file.

Studies conducted using the English language were included. Articles on overall UHC (UHC effective service coverage and FRP), UHC effective service coverage, UHC without specification with service coverage and FRP, and that reported quantitatively coverage, quality, inequity, FRP in the outcome of the study or qualitatively explored UHC research objectives were included. Types of study design included were quantitative, qualitative, mixed-research, and review types. The search was narrowed to include only literature published after 2015 in order to find studies that addressed the SDG target of 3.8 in 2015 and subsequent years. Articles were searched between October 20, 2021 and November 12, 2021, with the most recent update on March 03, 2022. Non-English language literature, abstracts only, comments or letters to the editor, erratum, corrections, and brief communications were all excluded.

### Study selection

In consultation with YA, AE developed and tested study selection forms on a random sample of references derived from search results. A second meeting was held to approve the study screening form and process. The inclusion and exclusion criteria were followed during the article screening process. Articles’ titles, abstracts, and full texts were reviewed in stages. After duplicates were removed using EndNote desktop x7 software and manual duplication removal, titles were screened. After that, abstracts were used to screen the literature. Those who passed abstract review were eligible for full-text review. Full-text articles were also screened for data charting purposes. For articles with only an abstract, contact was made with the study’s corresponding authors.

### Data charting

A piloted and refined data extraction tool was initially developed to chart the results of the review from full-text literature. Data was examined, charted, and sorted according to key issues and themes. Author(s), publication year, WHO geographic category, WB group, study approach, studied domain or topic, UHC themes, and health care service types were all extracted.

### Collation, summarization, and report of results

Based on years of publication, studied dimensions (interrelated objectives), WHO region, WB group, study approach, and health care service types, available articles were compiled and summarized with frequency and percentage.

A simple descriptive analysis was performed, and the results were presented in the form of tables and figures. The data reporting scheme was adjusted as needed based on the findings.

## Results

### Search results

PubMed (n = 6,230) and Web of Science (n = 832) databases were searched. Google Scholar (n=21), WB Library (n=5), WHO Library (n=7), UN Digital Library Collections (n=13), and Google (n=63) were also manually searched. A total of 7,171 records were discovered. Titles and abstracts were screened after duplicate records were removed both automatically and manually. Following that, 65 articles were chosen for full-text review. Finally, 47 articles were selected for scoping review. Figure depicts the PRISMA flow diagram of the study screening and selection process.

**Figure:**
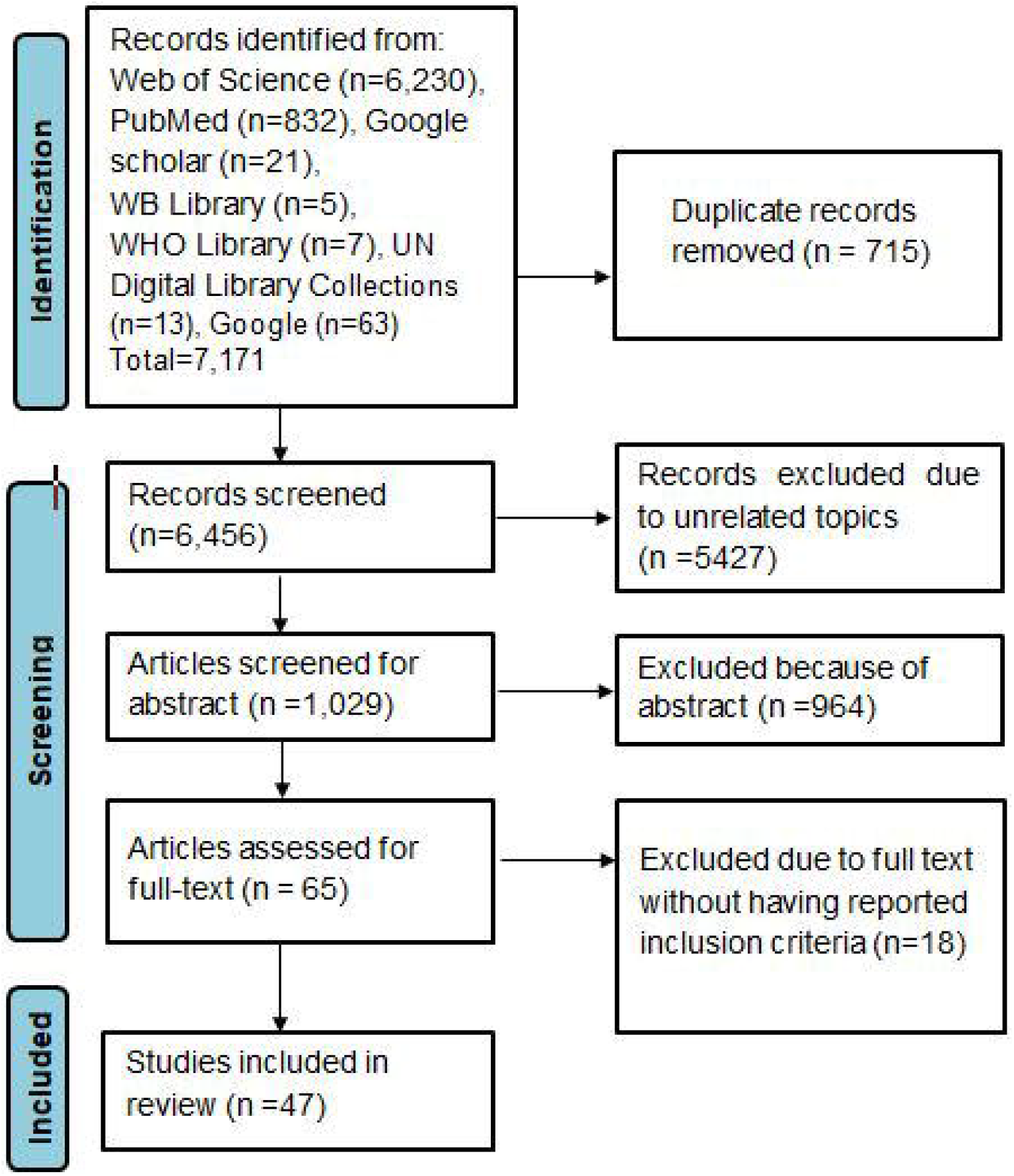
PRISMA-ScR flow diagram for articles selection process.

### Articles characteristics

Almost one-fourth of articles are from Africa and another 25.5% are across two or more WHO regions. According to income category, 42.6% are from lower-middle-income countries followed by 29.8% across two or more WB economy groups. More than half of the articles (54.1%) followed a quantitative research approach (Table).

**Table:**
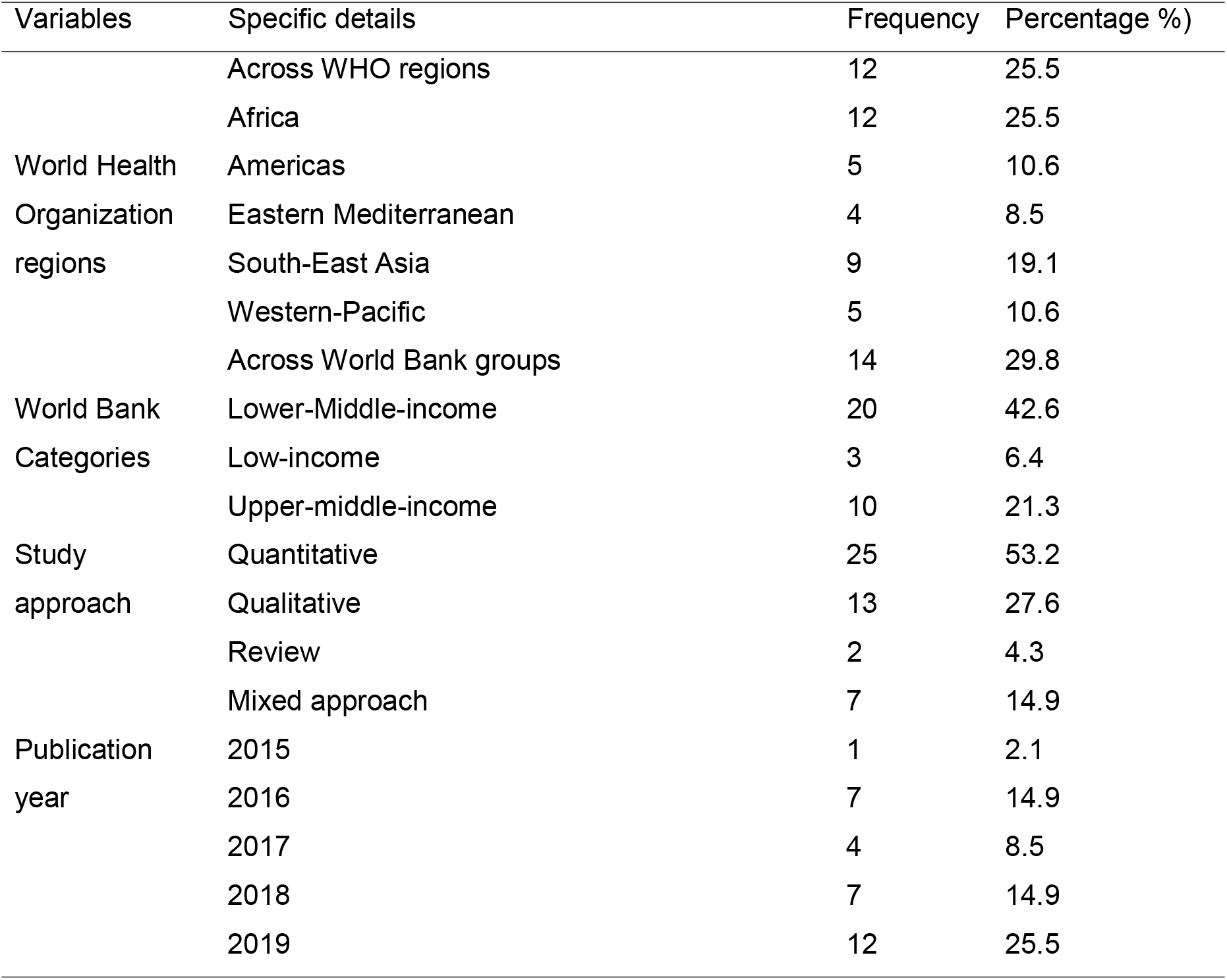
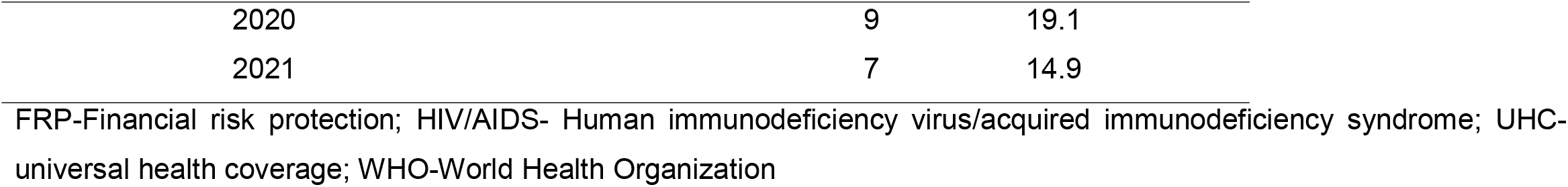
Articles distributions by World Health Organization region and World Bank category, study approach, interrelated objectives, health service type and year of study/publication (n=47)

| Variables | Specific details | Frequency | Percentage %) |
| --- | --- | --- | --- |
| World Health Organization regions | Across WHO regions | 12 | 25.5 |
|  | Africa | 12 | 25.5 |
|  | Americas | 5 | 10.6 |
|  | Eastern Mediterranean | 4 | 8.5 |
|  | South-East Asia | 9 | 19.1 |
|  | Western-Pacific | 5 | 10.6 |
| World Bank Categories | Across World Bank groups | 14 | 29.8 |
|  | Lower-Middle-income | 20 | 42.6 |
|  | Low-income | 3 | 6.4 |
|  | Upper-middle-income | 10 | 21.3 |
| Study approach | Quantitative | 25 | 53.2 |
|  | Qualitative | 13 | 27.6 |
|  | Review | 2 | 4.3 |
|  | Mixed approach | 7 | 14.9 |
| Publication year | 2015 | 1 | 2.1 |
|  | 2016 | 7 | 14.9 |
|  | 2017 | 4 | 8.5 |
|  | 2018 | 7 | 14.9 |
|  | 2019 | 12 | 25.5 |

|  |  |  |
| --- | --- | --- |
| 2020 | 9 | 19.1 |
| 2021 | 7 | 14.9 |

FRP-Financial risk protection; HIV/AIDS- Human immunodeficiency virus/acquired immunodeficiency syndrome; UHC- universal health coverage; WHO-World Health Organization

### Health Service types

Twenty articles (22-41) are categorized under health promotion, focusing on pathways, efforts, program evaluation and change, opportunities and challenges, barriers/factors/enablers, community-based health planning and service initiative, perceived effect of health reform on UHC, health-seeking behaviour and knowledge, health security and health promotion activities, the impact of insurance on coverage, and SCA dimensions of UHC. Health promotion encompasses funding and infrastructure, health literacy, the development of healthy public policies, the creation of supportive environments, and the strengthening of community actions and skills, as well as any activities that assist governments, communities, and individuals in dealing with and addressing health challenges.

Six articles discussed treatment aspects of health services, which were access to care for illness, access to treatment for rheumatic heart diseases, neglected tropical diseases (NTDs), mental disorders and hypertension (42-47).

According to GBD-2019, WHO and WB tracers, FP and/or SCA components for promotion, immunisation for prevention and other diseases in RMNC, IDs, NCDs for treatment aspects, nineteen articles were a combination of promotion, prevention, and treatment aspects (13, 48-65).

One study looked at the promotion and treatment of health services together (66).

One study on the quality of health care for disabled people (67) was classified as a palliative and rehabilitative care article, despite the fact that it did not adhere to palliative care assessment guidelines.

### Components and dimensions of UHC

The main four components of UHC are RMNC, IDs, NCDs and SCA. Distribution RMNC were reported in 40.4% of articles; NCDs in 36.2%, CDs in 25.5%, and SCA in 25.5%. Regarding dimensions, coverage was assessed by 53.2% of articles; FRP by 27.7%, inequity by 25.5% and quality by 6.4% of the articles (Supplementary file 3).

### Tracer indicators for summary measure of UHC

Of 25 quantitative articles, 19 articles used various tracer indicators to assess UHC quantitatively; the remaining six quantitative articles assessed each empirical analysis of the potential impact of importing health services, access and financial protection of emergency cares, perceived availability and quality of care, the performance of district health systems, crude coverage and financial protection, health-seeking behaviour and OOP health expenditures, the performance of health system.

The distribution of tracers in UHC effective service coverage estimation is described in the text below, based on WHO and WB, GBD-2019, and newly developed tracers. As indicator of RMNC, fourteen articles in the summary index of UHC effective service coverage included immunisations (13, 48, 51-53, 55-57, 59-65, 68); fifteen articles ANC and delivery (48, 49, 51-53, 55-57, 59-61, 63-65, 68), FP (13, 48, 51, 53, 55, 57, 59-64, 68) and child care seeking for pneumonia (13, 51, 52, 55-57, 59-61, 63-65, 68) each in thirteen articles. Other tracers used were under-5 diarrhoea treatment (13, 49, 52, 53, 55-57, 65, 68) and skilled birth attendance (48, 49, 51, 52, 55-57, 65, 68) each in nine articles, perinatal care for newborn babies and mothers (13, 49, 53, 68) in four articles, exclusive breastfeeding three articles (51, 57, 68), iron and folic Acid (≥100) in one article (53), and tetanus toxoid (53).

From the IDs, water and adequate sanitation were used in nine articles (48, 55, 57, 59-64, 68); tuberculosis effective treatment (13, 48, 59-64) and HIV ART (13, 48, 59-64) each in eight articles, insecticide-treated bed nets in five articles (51, 57, 59, 63, 64), perinatal care for newborn babies and mothers in two articles (13, 68) and tuberculosis case detection in one article (48).

NCDs tracer indicators were mean fasting blood glucose/ diabetes treatment (13, 48, 49, 59-64) and non-use of tobacco (48, 57, 59-64, 68) each in nine articles, non-raised blood pressure/hypertension treatment in eight articles (48, 49, 59-64), cervical cancer screening/treatment in seven articles (13, 49, 56, 59, 63-65), breast cancer screening/ treatment in four articles (13, 49, 56, 65), and uterine cancer treatment (13), colon and rectum cancer treatment (13), non-overweight (68), acute lymphoid leukaemia treatment (13), asthma treatment (13), epilepsy treatment (13), appendicitis treatment (13), paralytic ileus and intestinal obstruction treatment (13), ischemic heart disease treatment (13), stroke treatment (13), chronic kidney disease treatment (13), chronic obstructive pulmonary disease treatment (13), provision of treatment or advice on physical activity or diet (42) each in one article.

Regarding SCA, health worker density was included in seven articles (14, 59-64); hospital bed density (59-64) and health security (59-64) each in six articles, access to essential medicine in four articles (14, 59, 63, 64), inpatient admission (52, 65) and utilization of outpatient services (14, 42) each in two articles and maintenance of a stable health state after the last outpatient visit in one article (42).

As to FRP, catastrophic healthcare spending is used in eleven articles (14, 42, 48, 52, 53, 55-57, 62, 65, 68), impoverishment by out of pocket (OOP) healthcare spending is considered in five articles (48, 55, 57, 62, 68) and poverty gap due to OOP health spending used in one article (62).

Accessibility and affordability (14), as well as curative care and quality of care components (53), were developed as new tracers each in one article.

### Barriers/challenges of UHC

#### Policy level barrier

It is related to financing system (23, 26, 27, 29, 32-34), governance and leadership (22, 23, 29, 33, 36, 37), regulation and supervision mechanism (33, 34, 36), poverty (27, 34, 40), inequity (27, 32), insurance-related problems (27, 32), accreditation of facilities (26), narrowness of the benefit package (26), inadequate multi-sector collaborations (29), limited capacity to UHC policy implementation donor driven vertical programs (29), service preparedness (32), social infrastructure and social sustainability (23), failures in the expansion/shortage of services, technology and equipment and poor infrastructure (36, 47).

#### Health sector’s level barriers

Human resources shortage (33), deficient training (26, 33), low motivation (33), retention issues (33), skill-mix imbalance (33), health care quality problem (26, 33, 37), lack of guideline (33), political interference (33), ineffective or inadequate monitoring and supervision (27, 33), professional recruitment mechanisms (33), lack of attention to marginalized population (33), health infrastructure (26), inadequate number of health workers (26, 29, 36), service delivery (23), absence of diagnosis of the priority demands or conflict in setting priority (36, 38), behaviour(34), disease pattern (34), inadequate health system to early diagnose (47), and delayed reimbursement using prepaid health insurance (47) are barriers solved at health sector level.

#### Demand-side/ Cross cutting barrier

Sociocultural issues, historical mistrust to insurance prepayment, people perception that health care should be free of cost, lack of empowerment and information (33), health profile disparities between districts (37), unhealth life style (47), poor health seeking behaviour (47) and poor adherence to medication (47) can be demand sided barriers. Lack of common understanding on UHC and initiative (22, 33, 38), global movements (23), population health status (23), coverage (23), social determinants of health (37), emerging of non-communicable diseases (40) and inequities in access to health services or income (29, 37, 40) are cross-cutting barriers.

## Discussion

The purpose of this scoping review was to map existing research, identify gaps, and the most researched UHC dimensions, components and summarized main findings. Many articles were found in the African region and in countries with middle-income (lower and upper). The majority of the studies employed a quantitative research approach. Palliative and rehabilitative health care types did not well addressed in UHC research. The dimensions of service and financial coverage were most frequently studied, followed by inequity and quality of health care services

The current evidence found a greater number of articles than a scoping review of African implementation research of UHC (69). This is because the former was conducted on a single continent and concentrated on UHC research approaches. A bibliometric analysis, on the other hand, discovered a greater number of available evidence than the current scoping review (70). This is due to the fact that it includes all available evidence as terminology, title, phrases, or words in policy documents, commentaries, editorials, and all frequency counts found in databases by the first search without the conditions of pre-established exclusion criteria. Aside from that, the bibliometric analysis included articles dating back to 1990. UHC is a global agenda that has improved the health of the global population through political support, funding, and active national and international collaborations (71, 72). As a result, the number of research output is likely to increase over time, though current evidence shows that comparable numbers of articles are available each year. An earlier bibliometric analysis discovered an increasing trend in UHC research outputs (70).

Many of the studies in this review used a quantitative approach. A prior scoping review conducted in Africa discovered that qualitative and mixed-methods studies were the commonest method to investigate UHC (16). The former study did not take into account research on financial protection, UHC effective service and crude coverage, service capacity and access and HIV/AIDS treatment-related studies. UHC is intended to be quantified numerically as a summary index to track the progress of health care performance. Given the nature of UHC, appropriate articles used qualitative methods to investigate its challenges, opportunities, and success of UHC. Different health systems and policies in low, middle, and high-income countries may present different barriers and facilitators to attaining UHC (73-75).

The current study has revealed that the African region plays a leading role. The number of countries in the European region and the high-income category is large (76, 77). On the contrary, a substantial amount of research evidence was produced in middle-income countries. Trend analysis in health policy and systems research discovered an increasing trend of publications in low-and middle-income countries between 2003 and 2009 (78). This variation could be attributed to the nature of the health problems and the health policy in place regarding health research. Furthermore, even when contradictory findings are available, health research budgets and clinical trial infrastructures may determine health research activities in each continent. Evidence from a review finding of health science research in Africa indicated that nations with significant donor investment in health research may not necessarily produce a large number of health researchers (79). Articles were available across WHO regions that were comparable to African studies. UHC is a global strategy with agreed-upon tracers that aid in monitoring the global process towards universal access to health care. The available global reference framework for monitoring UHC is appropriate for conducting research at the multicounty level within a global context.

In 2019, the burden of NCDs was 63.8 percent worldwide, followed by CDs, RMNC, and nutritional disease (26.4 percent) (80). In the summary measure of the UHC index, RMNC was the most frequently studied component, followed by NCDs. This could be because many of the articles in the current review came from Africa and lower-and middle-income countries. In these countries, maternal and child morbidity and mortality are extremely high (81), making RMNC more likely to be investigated in UHC contexts. Similarly, a scoping review study on maternal, neonatal, and child health realized a high rate of publication in the most recent period (82). IDs and SCA had a lower frequency in the overall estimation of the UHC index based on the current review.

This review provides an answer to the question of how much UHC is universal and how much UHC is covered in the current health systems and policy research. UHC tracer indicators are focused on health promotion, disease prevention, treatment, palliative, and rehabilitative health care services at the individual and population level. Promotion aspects of health services were more frequently investigated in the current review. This could be due to the fact that those articles on the overall UHC that were non-specific to either component of UHC were more likely to be classified as belonging to health promotion. A single study was conducted on disabled population that was more likely to be close to palliative care health care services, though it was not in accordance with palliative care assessment protocol. Palliative care focuses on the physical, social, psychological, spiritual, and other issues confronting adults and children living with and dying from life-limiting conditions, as well as their families (83). Assessment of pain and symptom management, functional status, psychosocial care, caregiver assessment, and quality of life are all part of a palliative care measurement and evaluation tool (84). The Worldwide Hospice Palliative Care Alliance recommended conducting research to improve palliative care coverage (85) in order to ensure equitable health care access for the more than 40 million people worldwide who require palliative care each year (86). However, UHC effective service coverage measurement indicators are appropriate only for assessing the promotion, prevention and treatment aspects of health care, even though all health care services are theoretically expected to be covered globally (13).

In terms of dimensions, coverage was more commonly studied. The framework for monitoring and tracking was initially established for effective service coverage and FRP. UHC’s service coverage is a collection of many individual disease indicators that are used to assess the performance of the health care system as a whole. Therefore, it is not surprising that many articles have been written about the coverage dimension. Aside from the usual trend of calculating the service coverage summary index, a few articles calculated UHC by combining effective service coverage and FRP indicators. Although the effective coverage dimension expresses quality, intervention need and use together, there is clear assurance that it would address and measured comprehensively. In the current review, a few studies assessed the quality of care as a dimension of UHC; a single study developed a distinct quality of care measurement that was integrated into the UHC matrix because these services are theoretically integrated. Effective service coverage is predicated on the assumption that those in need receive high-quality health care services. However, having a high UHC index value does not imply that high-quality care is provided for each specific disease. For example in countries with high UHC index value (87), quality medical care services were found to be inadequate for patients with chronic diseases (88). Quality of health care can also be assessed using structure, process, and outcome indicators in the healthcare system (89).Therefore, generally, measuring the quality of care for specific diseases is helpful for stakeholders, clinicians, and health policymakers working on specific health problems (90).

The UHC summary index is useful for comparing the national and subnational progress of UHC. Inequity in UHC service coverage was reported in articles, with disparity across countries and geographical demarcation within a country. One of UHC’s primary functions is to promote health equity (91), and equity has been identified as a measurable component of UHC (92). It is linked to social determinants that should be monitored over time, across or within different settings and populations (93). Nonetheless, none of the UHC articles examined health disparities based on age, gender, race or ethnicity, residence, education level, or socioeconomic status. Similarly, range, absolute or relative difference, concentration index, and Gini coefficient were not used as equity measurement techniques in the current review.

## Strength and Limitation

This is the first scoping review of UHC, and it is accompanied by the most recent articles. Our review identified UHC literature in each category of health service type.

In terms of limitations, this review included only articles conducted in English; articles conducted in other languages may have been missed, and geographical representation of UHC articles may have been over or underestimated for regions. When considering UHC dimensions, they may have a different level of research articles discovered if another mapping review is done for specific disease types.

## Conclusions

More research is needed in settings where UHC has not been thoroughly investigated qualitatively and using a mixed-method approach, in addition to continuing quantitative monitoring and tracking of UHC status. According to the current review, future research should focus on the quality and equity of UHC health care services. Given that the distinct nature of UHC tracers may limit UHC’s articles on health promotion, prevention, and treatment aspects, palliative and rehabilitative care services require attention in the future research environment. For specific health problems, a scoping review may be required to identify research gaps in the dimension of UHC for that specific disease.

## Data Availability

All relevant data are within the manuscript and its supporting information files

## Abbreviations

ANC: Antenatal Care
AR: Antiretroviral Therapy
CHI: Catastrophic Health Expenditure
FP: Family Planning
GBD2019: Global Burden Disease 2019
HIV/AIDS: Human Immunodeficiency Virus/Acquired Immunodeficiency Syndrome
IDs: Infectious diseases
NCDs: Non-communicable diseases
RMNC: Reproductive, Maternal, Neonatal and Child
SCA: Service Capacity and Access
SDGs: Sustainable Development Goals
UN: United Nations
UHC: Universal Health Coverage
WB: World Bank
WHO: World Health Organization

## Acknowledgements

Not applicable

## Authors’ contributions

AE: conceptualization, identify research questions, built search strategy, literature searching, write manuscript draft and final report; YA: conceptualization, identify research questions, built search strategy, supervision and approve final manuscript; CFG: conceptualization, supervision approve final manuscript; FA: supervision and approve final manuscript.

## Competing interest

Authors declared no conflict of interest

## Funding

No fund receive to conduct this review

---

**Supplementary file 1:** Preferred reporting items for systematic reviews and meta-analyses extension for scoping reviews (PRISMA-ScR) checklist

| SECTION<br>TITLE | ITEM | PRISMA-ScR CHECKLIST ITEM | PAGE # |
| --- | --- | --- | --- |
| Title | 1 | Identify the report as a scoping review. | 1 |
| **ABSTRACT** |  |  |  |
| Structured summary | 2 | Provide a structured summary that includes (as applicable): background, objectives, eligibility criteria, sources of evidence, charting methods, results, and conclusions that relate to the review questions and objectives. | 2 |
| **INTRODUCTION** |  |  |  |
| Rationale | 3 | Describe the rationale for the review in the context of what is already known. Explain why the review questions/objectives lend themselves to a scoping review approach. | 4 and 5 |
| Objectives | 4 | Provide an explicit statement of the questions and objectives being addressed with reference to their key elements (e.g., population or participants, concepts, and context) or other relevant key elements used to conceptualize the review questions and/or objectives. | 5 |
| **METHODS** |  |  |  |
| Protocol and registration | 5 | Indicate whether a review protocol exists; state if and where it can be accessed and if available, provide registration information, including the registration number. | 6 |
| Eligibility criteria | 6 | Specify characteristics of the sources of evidence used as eligibility criteria (e.g., years considered, language, and publication status), and provide a rationale. | 6 |
| Information sources | 7 | Describe all information sources in the search (e.g., databases with dates of coverage and contact with authors to identify additional sources), as well as the date the most recent search was executed. | 6 |
| Search | 8 | Present the full electronic search strategy for at least one database, including any limits used, such that it could be repeated. | 6 |
| Selection of sources of evidence | 9 | State the process for selecting sources of evidence (i.e., screening and eligibility) included in the scoping review. | 7 |
| Data charting process | 10 | Describe the methods of charting data from the included sources of evidence (e.g., calibrated forms or forms that have been tested by the team before their use, and whether data charting was done independently or in duplicate) and any processes for obtaining and confirming data | 7 and 8 |

| SECTION | ITEM | PRISMA-ScR CHECKLIST ITEM | PAGE # |
| --- | --- | --- | --- |
|  |  | from investigators. |  |
| Data items | 11 | List and define all variables for which data were sought and any assumptions and simplifications made. | 7 |
| Critical appraisal of individual sources of evidence | 12 | If done, provide a rationale for conducting a critical appraisal of included sources of evidence; describe the methods used and how this information was used in any data synthesis (if appropriate). | Not applicable |
| Synthesis of results | 13 | Describe the methods of handling and summarizing the data that were charted. | 8 |
| **RESULTS** |  |  |  |
| Selection of sources of evidence | 14 | Give numbers of sources of evidence screened, assessed for eligibility, and included in the review, with reasons for exclusions at each stage, ideally using a flow diagram. | 8 (Figure) |
| Characteristics of sources of evidence | 15 | For each source of evidence, present characteristics for which data were charted and provide the citations. | 8 & 9 (Table) |
| Critical appraisal within sources of evidence | 16 | If done, present data on critical appraisal of included sources of evidence (see item 12). | Not applicable |
| Results of individual sources of evidence | 17 | For each included source of evidence, present the relevant data that were charted that relate to the review questions and objectives. | 10-11 |
| Synthesis of results | 18 | Summarize and/or present the charting results as they relate to the review questions and objectives. | 12-13 |
| **DISCUSSION** |  |  |  |
| Summary of evidence | 19 | Summarize the main results (including an overview of concepts, themes, and types of evidence available), link to the review questions and objectives, and consider the relevance to key groups. | 14-17 |
| Limitations | 20 | Discuss the limitations of the scoping review process. | 18 |
| Conclusions | 21 | Provide a general interpretation of the results with respect to the review questions and objectives, as well as potential implications and/or next steps. | 18 |
| **FUNDING** |  |  |  |
| Funding | 22 | Describe sources of funding for the included sources of evidence, as well as sources of funding for the scoping review. Describe the role of the funders of the scoping review. | 19 |

Source: Tricco AC, Lillie E, Zarin W, O'Brien KK, Colquhoun H, Levac D, et al. PRISMA Extension for Scoping Reviews (PRISMA-ScR): Checklist and Explanation. Ann Intern Med. 2018;169:467–473. doi: [10.7326/M18-0850](https://doi.org/10.7326/M18-0850).

**Supplementary file 2:** Search strategy

| Database | Search strategy | Result | Filters applied: |
| --- | --- | --- | --- |
| PubMed | (Universal[Title/Abstract] AND (health[Title/Abstract] OR | 6,230 | From 2015/1/1 - |
|  | "health care"[Title/Abstract] OR healthcare[Title/Abstract] OR "health service"[Title/Abstract])) AND (access[Title/Abstract] OR coverage[Title/Abstract] OR Equity[Title/Abstract] OR disparity[Title/Abstract] OR inequity[Title/Abstract] OR equality[Title/Abstract] OR inequality[Title/Abstract] OR quality[Title/Abstract] OR expenditure [Title/Abstract] OR cost [Title/Abstract])<br>*Filters applied:* English, Humans, from 2015/1/1 - 2022/3/3. |  | 2022/3/3 |
| Web of Science | (TI=("Universal" ) AND TI=("health" OR "health care" OR "healthcare" OR "health service") AND TI=(access OR coverage OR equity OR disparity OR inequity OR equality OR inequality OR quality OR expenditure OR cost)) AND (PY==("2021" OR "2020" OR "2019" OR "2018" OR "2016" OR "2015" OR "2017")) AND English (Languages) Not Document type: Letters or Meeting Abstracts or Editorial Materials or Corrections or Book chapters or Books Or Book reviews or Biographical-Items | 832 | From 2015/1/1 - 2022/3/3 |

**Supplementary file 3:**
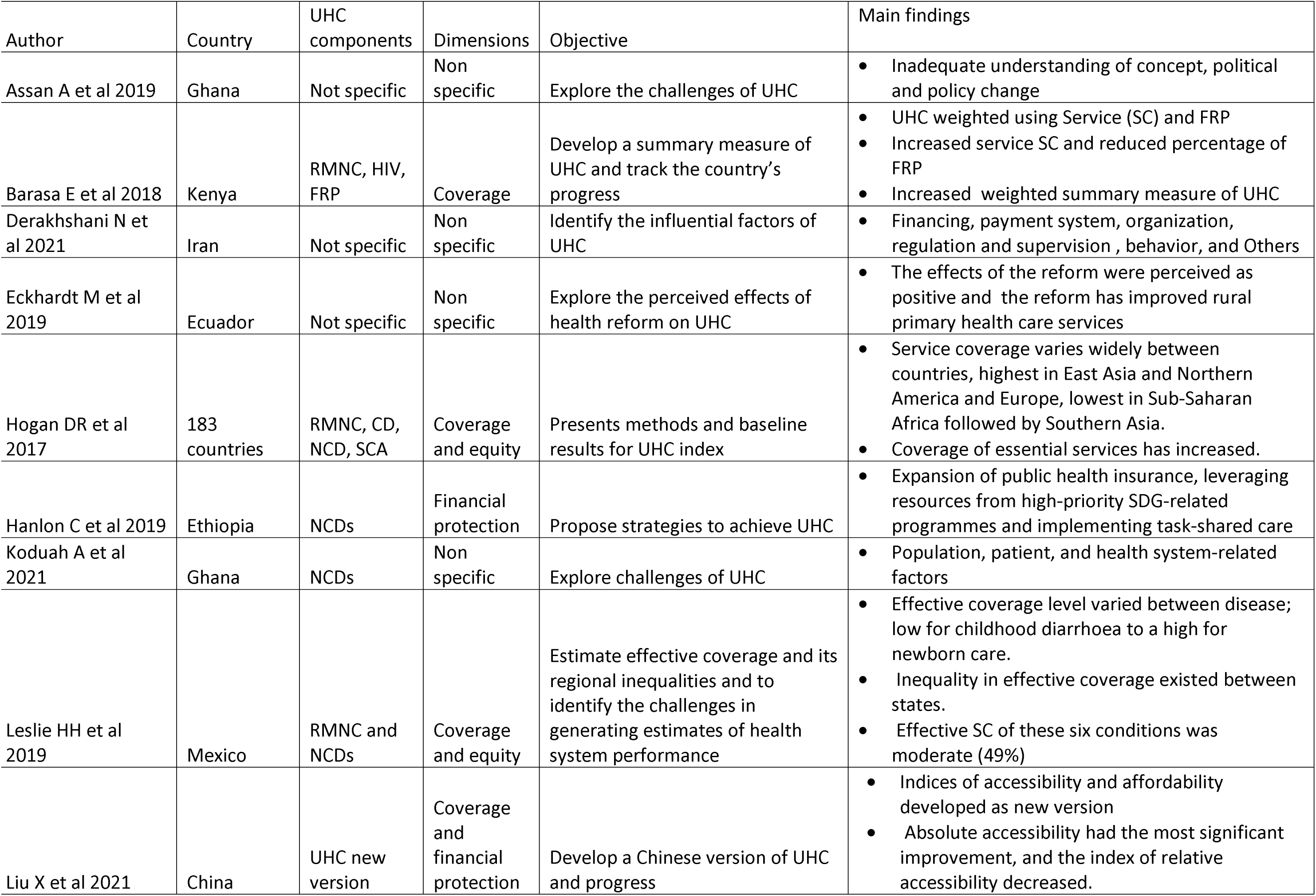
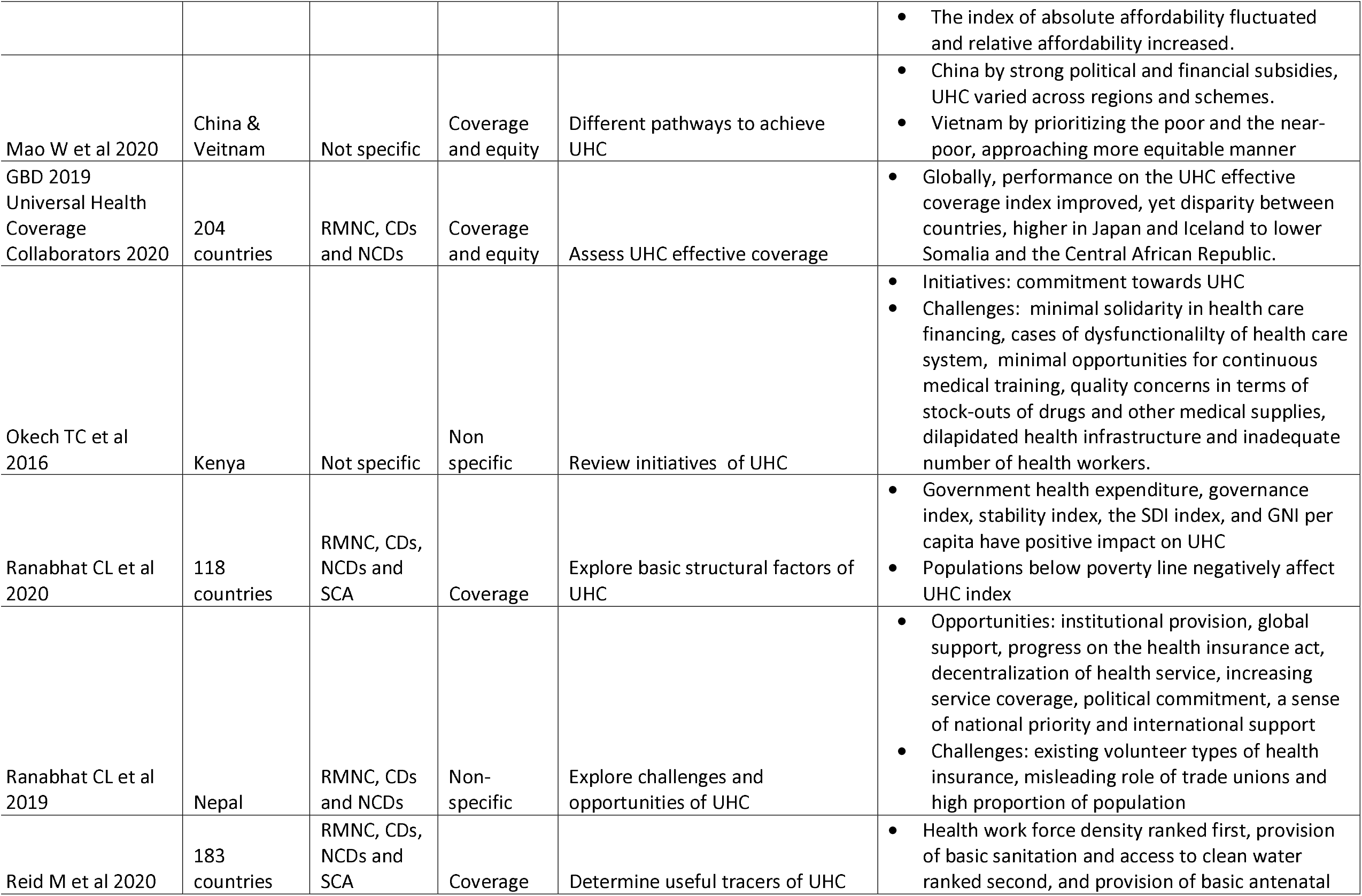
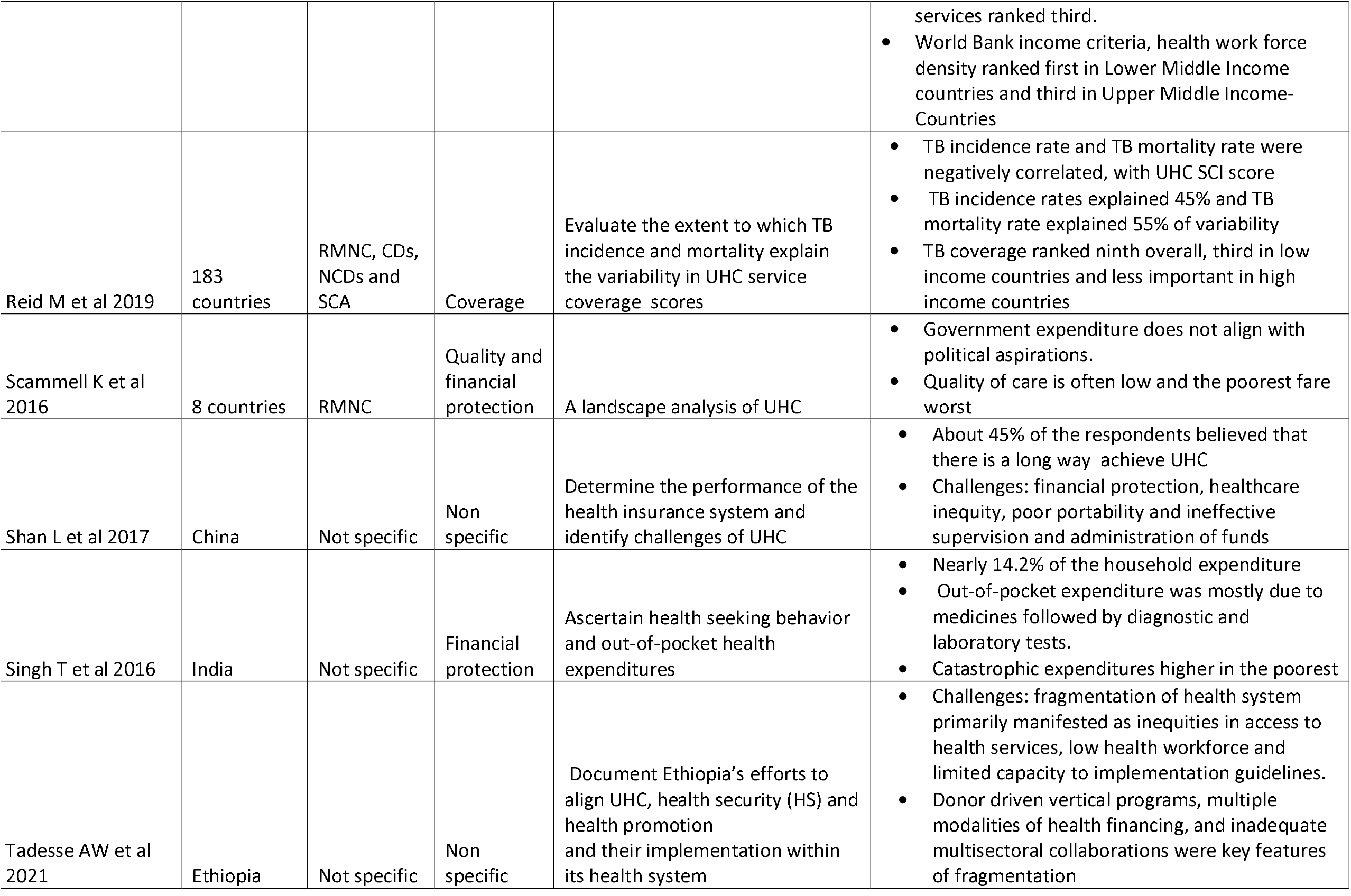
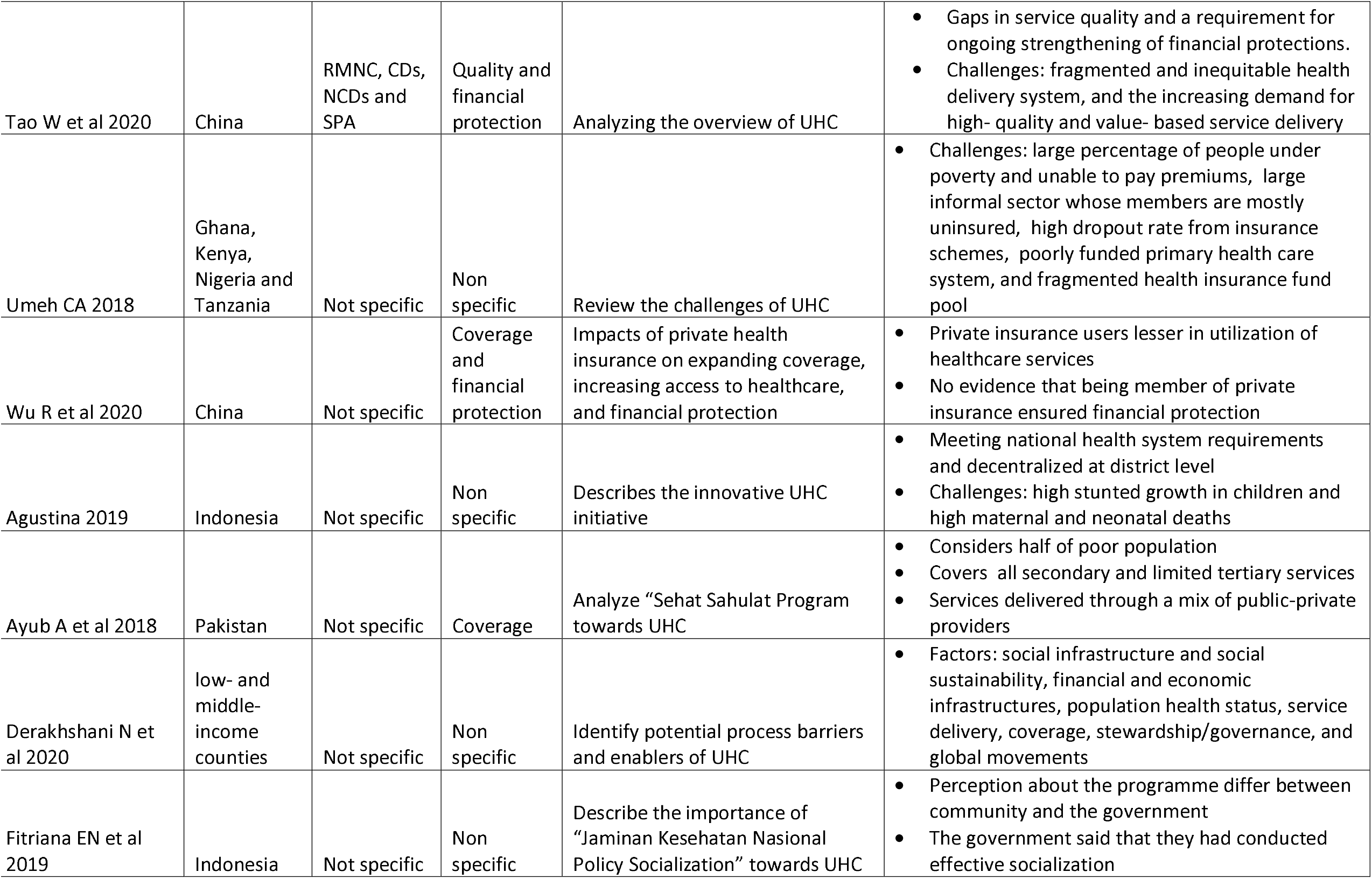
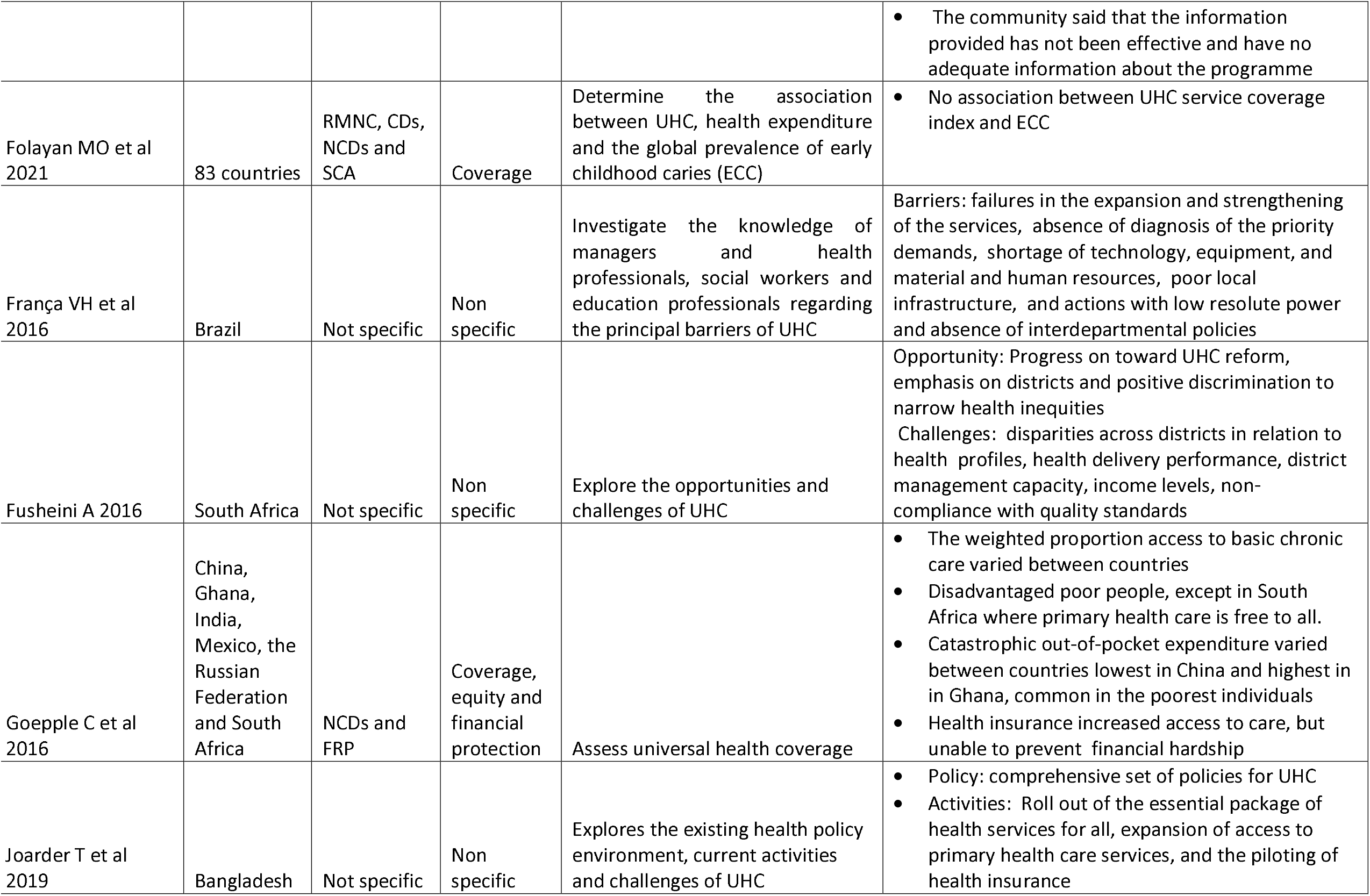
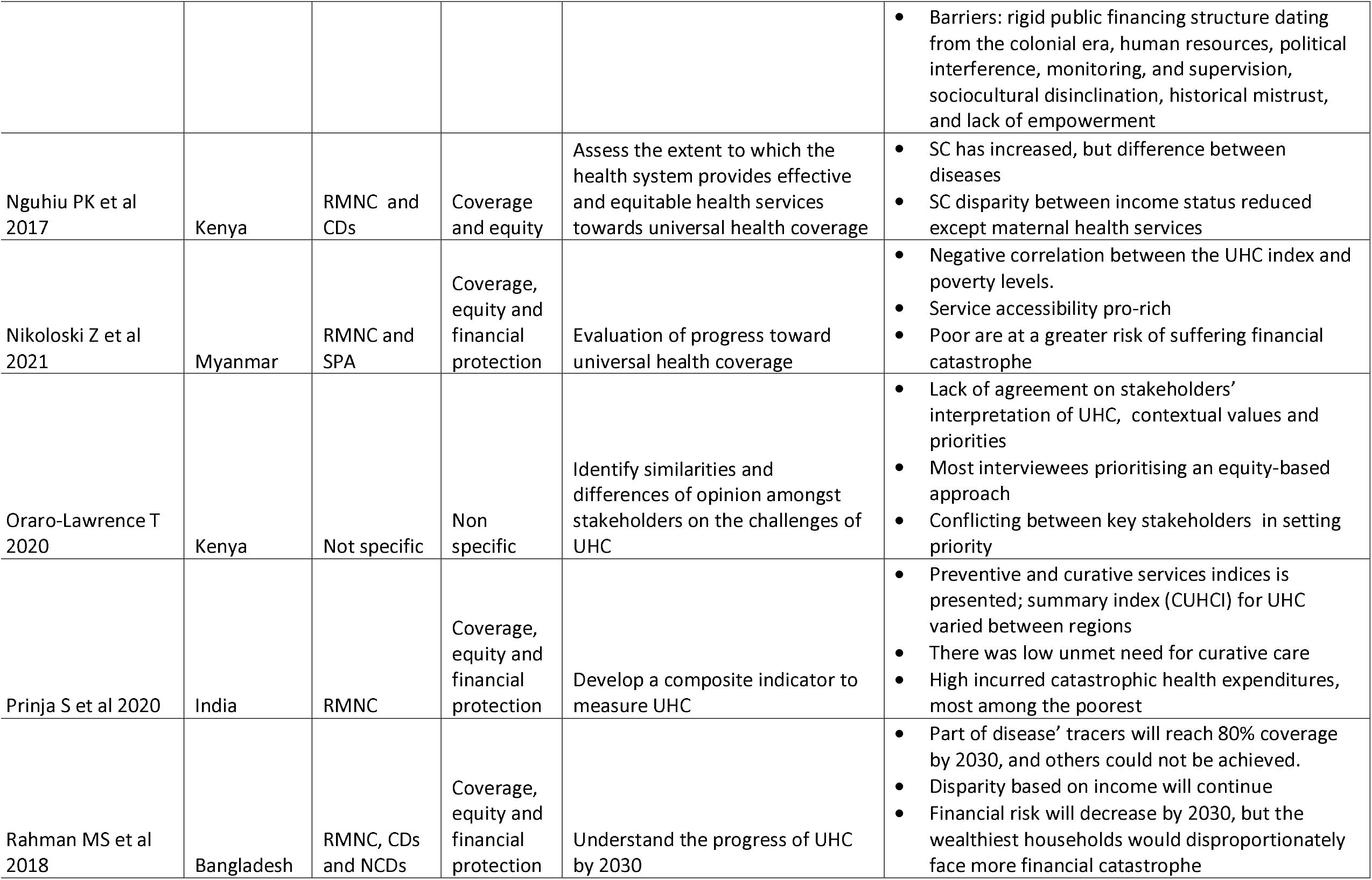
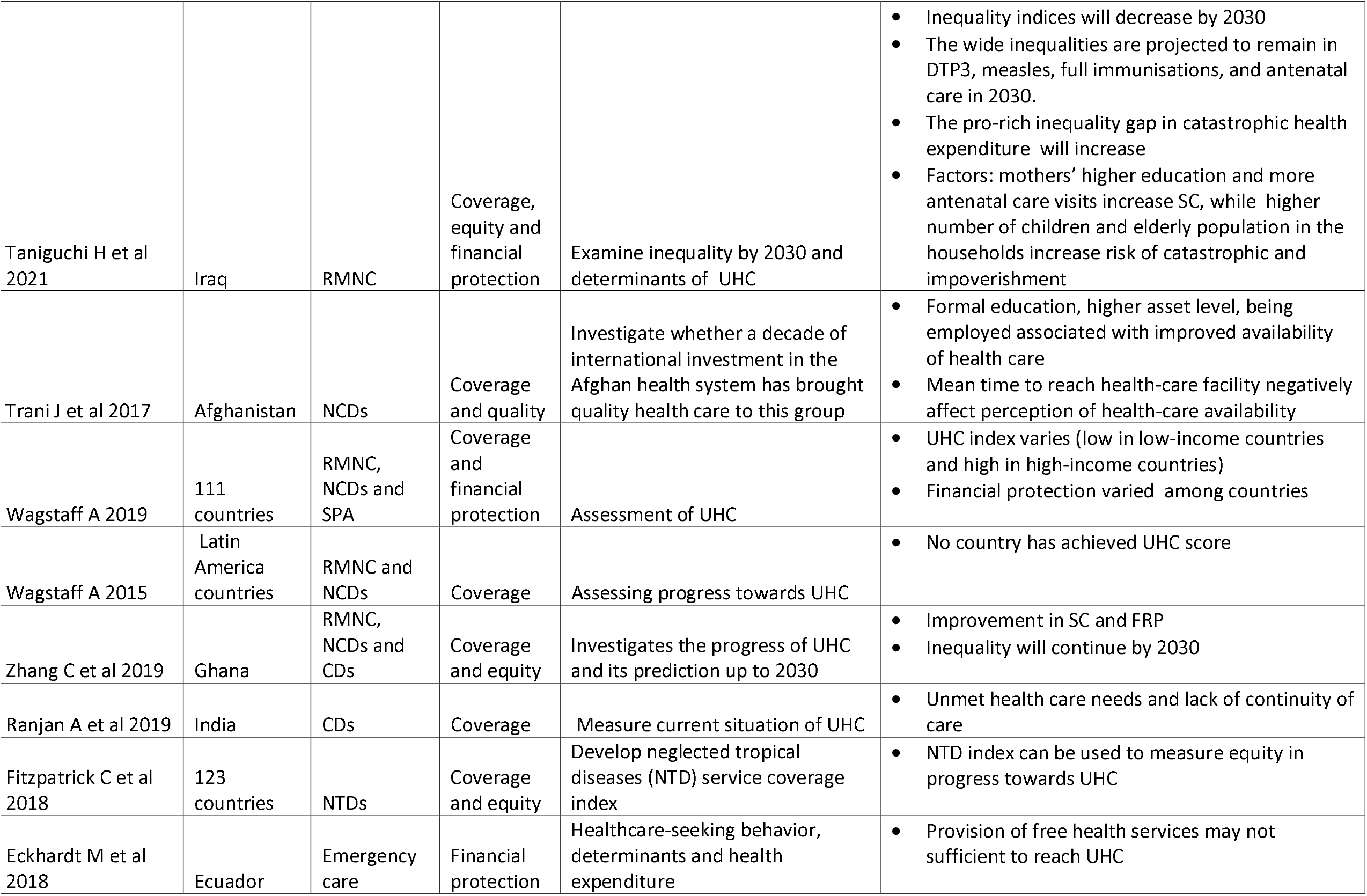
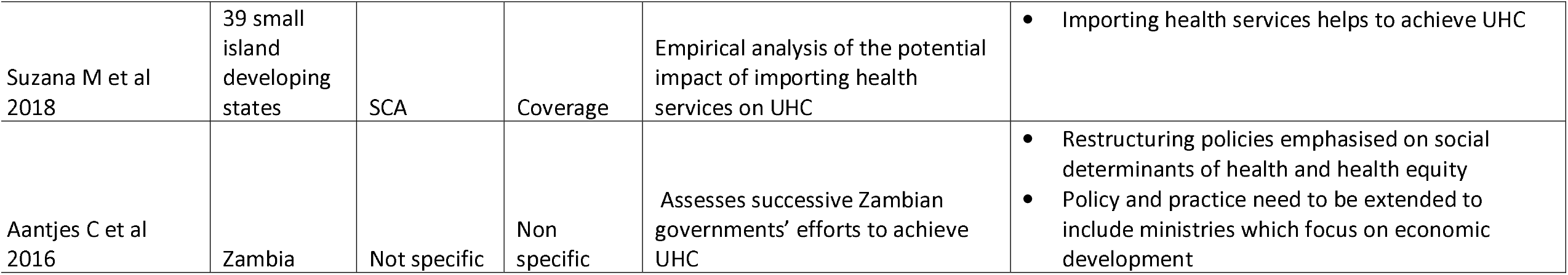
Articles’ main findings

## Notes

### Competing Interest Statement

The authors have declared no competing interest.

### Funding Statement

Authors did not receive fund

## References

1. Abiiro GA, De Allegri M. Universal health coverage from multiple perspectives: a synthesis of conceptual literature and global debates. Bmc International Health and Human Rights. 2015;15.

2. Thomson S, Cylus J, Evetovits T, editors. How you measure matters: monitoring financial protection to generate policy-relevant evidence for universal health coverage–lessons from Europe. Organized session 9 July 2017. IHEA 12th World Congress in Health Economics; 2017.

3. Tangcharoensathien V, Mills A, Palu T. Accelerating health equity: the key role of universal health coverage in the Sustainable Development Goals. Bmc Medicine. 2015;13.

4. UNDP. Universal health coverage for sustainable development - issue brief 2019 [cited 2021 October 2]. Available from: https://www.undp.org/publications/universal-health-coverage-sustainable-development-issue-brief.

5. World Health Organization. Declaration of alma-ata. World Health Organization. Regional Office for Europe, 1978.

6. United Nations. Resolution adopted by the General Assembly on 25 September 2015 2015 [cited 2021 23 October]. Available from: https://undocs.org/A/RES/70/1.

7. uhc 2030. State of commitment to universal health coverage: synthesis, 2020 2020 [cited 2021 October 2]. Available from: https://www.uhc2030.org/blog-news-events/uhc2030-news/state-of-commitment-to-universal-health-coverage-synthesis-2020-555434/.

8. United Nations. Universal Health Coverage: Moving Together to Build a Healthier World New York 2019 [cited 2021 24 October]. Available from: https://www.un.org/pga/73/event/universal-health-coverage/.

9. World Health Organization. The world health report: health systems financing: the path to universal coverage: executive summary. World Health Organization, 2010.

10. World Health Organization. Research for universal health coverage: World health report 2013 2013 [cited 2021 15 August]. Available from: https://www.who.int/publications/i/item/9789240690837.

11. Kim JY. Speech by World Bank Group President Jim Yong Kim at the Government of Japan-World Bank Conference on universal health coverage. The World Bank, 2013.

12. World Health Organization. Universal health coverage (UHC) 2021, April 1. Available from: https://www.who.int/news-room/fact-sheets/detail/universal-health-coverage-(uhc).

13. Lozano R, Fullman N, Mumford JE, Knight M, Barthelemy CM, Abbafati C, et al. Measuring universal health coverage based on an index of effective coverage of health services in 204 countries and territories, 1990–2019: a systematic analysis for the Global Burden of Disease Study 2019. The Lancet. 2020;396(10258):1250–84.

14. Liu X, Wang Z, Zhang H, Meng Q. Measuring and evaluating progress towards Universal Health Coverage in China. Journal of global health. 2021;11.

15. Daudt HM, van Mossel C, Scott SJ. Enhancing the scoping study methodology: a large, inter-professional team’s experience with Arksey and O’Malley’s framework. BMC medical research methodology. 2013;13(1):1–9.

16. Nnaji CA. Implementation research approaches to promoting universal health coverage in Africa: a scoping review. The International journal of health planning and management. 2021;21(1):414.

17. Moreno-Betancur M, Latouche A, Menvielle G, Kunst AE, Rey G. Relative index of inequality and slope index of inequality: a structured regression framework for estimation. Epidemiology. 2015;26(4):518–27.

18. Arksey H, O’Malley L. Scoping studies: towards a methodological framework. International journal of social research methodology. 2005;8(1):19–32.

19. Levac D, Colquhoun H, O’Brien KK. Scoping studies: advancing the methodology. Implementation science. 2010;5(1):1–9.

20. Peters MDJ, Marnie C, Tricco AC, Pollock D, Munn Z, Alexander L, et al. Updated methodological guidance for the conduct of scoping reviews. JBI evidence implementation. 2021;19(1):3–10.

21. Tricco AC, Lillie E, Zarin W, O’Brien KK, Colquhoun H, Levac D, et al. PRISMA Extension for Scoping Reviews (PRISMA-ScR): Checklist and Explanation. Ann Intern Med. 2018;169(7):467–73.

22. Assan A, Takian A, Aikins M, Akbarisari A. Challenges to achieving universal health coverage through community-based health planning and services delivery approach: a qualitative study in Ghana. BMJ open. 2019;9(2):e024845.

23. Derakhshani N, Doshmangir L, Ahmadi A, Fakhri A, Sadeghi-Bazargani H, Gordeev VS. Monitoring process barriers and enablers towards universal health coverage within the sustainable development goals: a systematic review and content analysis. ClinicoEconomics and Outcomes Research: CEOR. 2020;12:459.

24. Eckhardt M, Carlfjord S, Faresjo T, Crespo-Burgos A, Forsberg BC, Falk M. Universal Health Coverage in Marginalized Populations: A Qualitative Evaluation of a Health Reform Implementation in Rural Ecuador. Inquiry-the Journal of Health Care Organization Provision and Financing. 2019;56.

25. Mao W, Tang Y, Tran T, Pender M, Khanh PN, Tang S. Advancing universal health coverage in China and Vietnam: lessons for other countries. BMC Public Health. 2020;20(1):1–9.

26. Okech TC, Lelegwe SL. Analysis of universal health coverage and equity on health care in Kenya. Global journal of health science. 2016;8(7):218.

27. Shan L, Wu Q, Liu C, Li Y, Cui Y, Liang Z, et al. Perceived challenges to achieving universal health coverage: a cross-sectional survey of social health insurance managers/administrators in China. BMJ open. 2017;7(5):e014425.

28. Singh T, Roy P, Jamir L, Gupta S, Kaur N, Jain D, et al. Assessment of universal healthcare coverage in a district of North India: a rapid cross-sectional survey using tablet computers. PLoS One. 2016;11(6):e0157831.

29. Tadesse AW, Gurmu KK, Kebede ST, Habtemariam MK. Analyzing efforts to synergize the global health agenda of universal health coverage, health security and health promotion: a case-study from Ethiopia. Globalization and Health. 2021;17(1):1–13.

30. Umeh CA. Challenges toward achieving universal health coverage in Ghana, Kenya, Nigeria, and Tanzania. The International journal of health planning and management. 2018;33(4):794–805.

31. Wu R, Li N, Ercia A. The effects of private health insurance on universal health coverage objectives in China: a systematic literature review. International Journal of Environmental Research and Public Health. 2020;17(6):2049.

32. Agustina R, Dartanto T, Sitompul R, Susiloretni KA, Achadi EL, Taher A, et al. Universal health coverage in Indonesia: concept, progress, and challenges. The Lancet. 2019;393(10166):75–102.

33. Ayub A, Khan RS, Khan SA, Hussain H, Tabassum A, Shehzad JA, et al. Progress of Khyber Pakhtunkhwa (Pakistan) towards universal health coverage. Journal of Ayub Medical College Abbottabad. 2018;30(3):481–4.

34. Derakhshani N, Maleki M, Pourasghari H, Azami-Aghdash S. The influential factors for achieving universal health coverage in Iran: a multimethod study. BMC health services research. 2021;21(1):1–13.

35. Fitriana EN, Probandari AN, Pamungkasari EP, Ardyanto TD, Puspitaningrum RA. The importance of socialization in achieving universal health coverage: Case study of Jaminan Kesehatan Nasional (JKN) implementation in two different region in Central Java province. JKKI: Jurnal Kedokteran dan Kesehatan Indonesia. 2019;10(2):110–20.

36. França VHd, Modena CM, Confalonieri UEC. A multiprofessional perspective on the principal barriers to universal health coverage and universal access to health in extremely poor territories: the contributions of nursing. Revista Latino-Americana de Enfermagem. 2016;24.

37. Fusheini A, Eyles J. Achieving universal health coverage in South Africa through a district health system approach: conflicting ideologies of health care provision. BMC Health Services Research. 2016;16(1):1–11.

38. Oraro-Lawrence T, Wyss K. Policy levers and priority-setting in universal health coverage: a qualitative analysis of healthcare financing agenda setting in Kenya. BMC health services research. 2020;20(1):1–11.

39. Suzana M, Walls H, Smith R, Hanefeld J. Achieving universal health coverage in small island states: could importing health services provide a solution? BMJ global health. 2018;3(1):e000612.

40. Aantjes C, Quinlan T, Bunders J. Towards universal health coverage in Zambia: impediments and opportunities. Development in Practice. 2016;26(3):298–307.

41. Joarder T, Chaudhury TZ, Mannan I. Universal health coverage in Bangladesh: activities, challenges, and suggestions. Advances in Public Health. 2019;2019.

42. Goeppel C, Frenz P, Grabenhenrich L, Keil T, Tinnemann P. Assessment of universal health coverage for adults aged 50 years or older with chronic illness in six middle-income countries. Bulletin of the World Health Organization. 2016;94(4):276.

43. Ranjan A, Thiagarajan S, Garg S, Danda D. Progress towards universal health coverage in the context of rheumatic diseases in India. International Journal of Rheumatic Diseases. 2019;22(5):880–9.

44. Fitzpatrick C, Bangert M, Mbabazi PS, Mikhailov A, Zouré H, Rebollo MP, et al. Monitoring equity in universal health coverage with essential services for neglected tropical diseases: an analysis of data reported for five diseases in 123 countries over 9 years. The Lancet Global Health. 2018;6(9):e980–e8.

45. Eckhardt M, Santillán D, Faresjö T, Forsberg BC, Falk M. Universal health coverage in rural Ecuador: a cross-sectional study of perceived emergencies. Western Journal of Emergency Medicine. 2018;19(5):889.

46. Hanlon C, Alem A, Lund C, Hailemariam D, Assefa E, Giorgis TW, et al. Moving towards universal health coverage for mental disorders in Ethiopia. International journal of mental health systems. 2019;13(1):1–16.

47. Koduah A, Nonvignon J, Colson A, Kurdi A. Health systems, population and patient challenges for achieving universal health coverage for hypertension in Ghana. 2021;36(9):1451–8.

48. Barasa E, Nguhiu P, McIntyre D. Measuring progress towards sustainable development goal 3.8 on universal health coverage in Kenya. BMJ global health. 2018;3(3):e000904.

49. Leslie HH, Doubova SV, Pérez-Cuevas R. Assessing health system performance: effective coverage at the Mexican Institute of social security. Health Policy and Planning. 2019;34(Supplement_2):ii67–ii76.

50. Scammell K, Noble DJ, Rasanathan K, O’Connell T, Ahmed AS, Begkoyian G, et al. A landscape analysis of universal health coverage for mothers and children in South Asia. BMJ global health. 2016;1(1):e000017.

51. Nguhiu PK, Barasa EW, Chuma J. Determining the effective coverage of maternal and child health services in Kenya, using demographic and health survey data sets: tracking progress towards universal health coverage. Tropical Medicine & International Health. 2017;22(4):442–53.

52. Nikoloski Z, McGuire A, Mossialos E. Evaluation of progress toward universal health coverage in Myanmar: A national and subnational analysis. PLoS medicine. 2021;18(10):e1003811.

53. Prinja S, Gupta R, Bahuguna P, Sharma A, Kumar Aggarwal A, Phogat A, et al. A composite indicator to measure universal health care coverage in India: way forward for post-2015 health system performance monitoring framework. Health policy and planning. 2017;32(1):43–56.

54. Rahman MS, Rahman MM, Gilmour S, Swe KT, Abe SK, Shibuya K. Trends in, and projections of, indicators of universal health coverage in Bangladesh, 1995-2030: a Bayesian analysis of population-based household data. Lancet Global Health. 2018;6(1):E84–E94.

55. Taniguchi H, Rahman MM, Swe KT, Islam MR, Rahman MS, Parsell N, et al. Equity and determinants in universal health coverage indicators in Iraq, 2000–2030: a national and subnational study. International journal for equity in health. 2021;20(1):1–10.

56. Wagstaff A, Dmytraczenko T, Almeida G, Buisman L, Hoang-Vu Eozenou P, Bredenkamp C, et al. Assessing Latin America’s progress toward achieving universal health coverage. Health Affairs. 2015;34(10):1704–12.

57. Zhang C, Rahman MS, Rahman MM, Yawson AE, Shibuya K. Trends and projections of universal health coverage indicators in Ghana, 1995-2030: A national and subnational study. PloS one. 2019;14(5):e0209126.

58. Ranabhat CL, Kim CB, Singh A, Acharya D, Pathak K, Sharma B, et al. Challenges and opportunities towards the road of universal health coverage (UHC) in Nepal: a systematic review. Arch Public Health. 2019;77:5.

59. Ranabhat CL, Jakovljevic M, Dhimal M, Kim C-B. Structural factors responsible for universal health coverage in low-and middle-income countries: results from 118 countries. Frontiers in public health. 2020:414.

60. Reid M, Gupta R, Roberts G, Goosby E, Wesson P. Achieving Universal Health Coverage (UHC): Dominance analysis across 183 countries highlights importance of strengthening health workforce. PloS one. 2020;15(3):e0229666.

61. Reid M, Roberts G, Goosby E, Wesson P. Monitoring Universal Health Coverage (UHC) in high Tuberculosis burden countries: Tuberculosis mortality an important tracer of UHC service coverage. PLoS One. 2019;14(10):e0223559.

62. Tao W, Zeng Z, Dang H, Li P, Chuong L, Yue D, et al. Towards universal health coverage: achievements and challenges of 10 years of healthcare reform in China. BMJ global health. 2020;5(3):e002087.

63. Hogan DR, Stevens GA, Hosseinpoor AR, Boerma T. Monitoring universal health coverage within the Sustainable Development Goals: development and baseline data for an index of essential health services. The Lancet Global Health. 2018;6(2):e152–e68.

64. Folayan MO, Tantawi ME, Virtanen JI, Feldens CA, Rashwan M, Kemoli AM, et al. An ecological study on the association between universal health service coverage index, health expenditures, and early childhood caries. BMC oral health. 2021;21(1):1–7.

65. Wagstaff A, Neelsen S. A comprehensive assessment of universal health coverage in 111 countries: a retrospective observational study. The Lancet Global Health. 2020;8(1):e39–e49.

66. Liu XY, Wang ZY, Zhang H, Meng QY. Measuring and evaluating progress towards Universal Health Coverage in China. Journal of global health. 2021;11.

67. Trani JF, Kumar P, Ballard E, Chandola T. Assessment of progress towards universal health coverage for people with disabilities in Afghanistan: a multilevel analysis of repeated cross-sectional surveys. Lancet Global Health. 2017;5(8):E828–E37.

68. Rahman MS, Rahman MM, Gilmour S, Swe KT, Abe SK, Shibuya K. Trends in, and projections of, indicators of universal health coverage in Bangladesh, 1995–2030: a Bayesian analysis of population-based household data. The Lancet Global Health. 2018;6(1):e84–e94.

69. Makadzange K, Radebe Z, Maseko N, Lukhele V, Masuku S, Fakudze G, et al. Implementation of Urban Health Equity Assessment and Response Tool: a Case of Matsapha, Swaziland. Journal of urban health : bulletin of the New York Academy of Medicine. 2018;95(5):672–81.

70. Ghanbari MK, Behzadifar M, Doshmangir L, Martini M, Bakhtiari A, Alikhani M, et al. Mapping research trends of universal health coverage from 1990 to 2019: Bibliometric analysis. JMIR Public Health and Surveillance. 2021;7(1):e24569.

71. Hammonds R, Ooms G, Mulumba M, Maleche A. UHC2030’s Contributions to Global Health Governance that Advance the Right to Health Care: A Preliminary Assessment. Health and Human Rights. 2019;21(2):235.

72. Ssengooba F, Ssennyonjo A, Rutebemberwa E, Musila T, Namusoke Kiwanuka S, Kemari E, et al. Research for universal health coverage: setting priorities for policy and systems research in Uganda. Global health action. 2021;14(1):1956752.

73. Saif-Ur-Rahman K, Mamun R, Nowrin I, Hossain S, Islam K, Rumman T, et al. Primary healthcare policy and governance in low-income and middle-income countries: an evidence gap map. BMJ global health. 2019;4(Suppl 8):e001453.

74. Mills A. Health care systems in low-and middle-income countries. New England Journal of Medicine. 2014;370(6):552–7.

75. Polin K, Hjortland M, Maresso A, van Ginneken E, Busse R, Quentin W. “Top-Three” health reforms in 31 high-income countries in 2018 and 2019: an expert informed overview. Health Policy. 2021;125(7):815–32.

76. Ajayi AI, Akpan W. Determinants of condom use among parous women in North Central and South Western Nigeria: a cross-sectional survey. BMC research notes. 2018;11(1):1–6.

77. World Health Organization. Countries 2022 [cited 2022 03]. Available from: https://www.who.int/countries.

78. Adam T, Ahmad S, Bigdeli M, Ghaffar A, Røttingen J-A. Trends in health policy and systems research over the past decade: still too little capacity in low-income countries. PloS one. 2011;6(11):e27263.

79. Wenham C, Wouters O, Jones C, Juma PA, Mijumbi-Deve RM, Sobngwi-Tambekou JL, et al. Measuring health science research and development in Africa: mapping the available data. Health Research Policy and Systems. 2021;19(1):1–13.

80. Furuoka F, Hoque MZ. Determinants of antiretroviral therapy coverage in Sub-Saharan Africa. PeerJ. 2015;3:e1496.

81. Paulson KR, Kamath AM, Alam T, Bienhoff K, Abady GG, Abbas J, et al. Global, regional, and national progress towards Sustainable Development Goal 3.2 for neonatal and child health: all-cause and cause-specific mortality findings from the Global Burden of Disease Study 2019. The Lancet. 2021;398(10303):870–905.

82. Chan GJ, Daniel J, Getnet M, Kennedy M, Olowojesiku R, Hunegnaw BM, et al. Gaps in maternal, newborn, and child health research: a scoping review of 72 years in Ethiopia. Newborn, and Child Health Research: A Scoping Review of. 2021;72.

83. World Health Organization. WHO launches global effort to halve medication-related errors in 5 years. Geneva/Bonn Available online: http://www.whoint/mediacentre/news/releases/2017/medication-related-errors/en/(accessed on 08 August 2019). 2017.

84. National Palliative Care Research Center. Measurement and Evaluation Tools 2013 [cited 2022 March 03]. Available from: http://www.npcrc.org/content/25/Measurement-and-Evaluation-Tools.aspx.

85. Morris C. Universal health coverage and palliative care. Do not Leave Those Suffering Behind London: World Hospice and Palliative Care Alliance. 2014.

86. Health Organization World. Palliative care Genieva5 August 2020 [cited 2021 25 January]. Available from: https://www.who.int/news-room/fact-sheets/detail/palliative-care.

87. Lozano R, Fullman N, Mumford JE, Knight M, Barthelemy CM, Abbafati C, et al. Measuring universal health coverage based on an index of effective coverage of health services in 204 countries and territories, 1990–2019: a systematic analysis for the Global Burden of Disease Study 2019. The Lancet. 2020;396(10258):1250–84.

88. Adé A, Debroucker F, Delporte L, De Monclin C, Fayet E, Legendre P, et al. Chronic patients’ satisfaction and priorities regarding medical care, information and services and quality of life: a French online patient community survey. BMC health services research. 2020;20(1):1–10.

89. Titi-Ofei R, Osei-Afriyie D, Karamagi H. Monitoring Quality of Care in the WHO Africa Region-A study design for measurement and tracking, towards UHC attainment. Global health action. 2021;14(1):1939493.

90. Gimeno-García A, Gimeno-García A, Franco-Moreno A, Montero-Hernández C, Montero-Hernández C, Arponen S, et al. Analysis of adherence to HIV-positive quality of care indicators and their impact on service quality perceptions in patients: A Spanish cross-sectional study. Health and Quality of Life Outcomes. 2020;18(1).

91. Tangcharoensathien V, Mills A, Palu T. Accelerating health equity: the key role of universal health coverage in the Sustainable Development Goals. BMC medicine. 2015;13(1):1–5.

92. Rodney AM, Hill PS. Achieving equity within universal health coverage: A narrative review of progress and resources for measuring success. International Journal for Equity in Health. 2014;13(1).

93. Dover DC, Belon AP. The health equity measurement framework: a comprehensive model to measure social inequities in health. International Journal for Equity in Health. 2019;18(1):1–12.

